# Narrowing gap in regional and age-specific excess mortality in the first year and a half of COVID-19 in Hungary

**DOI:** 10.1101/2022.01.05.22268786

**Authors:** Csaba G. Tóth

## Abstract

In the first year and a half of the pandemic, the excess mortality in Hungary was 28,400, which was 1,700 lower than the official statistics on COVID-19 deaths. This discrepancy can be partly explained by protective measures instated during the COVID-19 pandemic that decreased the intensity of the seasonal flu outbreak, which caused on average 3,000 deaths per year. Compared to the second wave of the COVID-19 pandemic, the third wave showed a reduction in the differences in excess mortality between age groups and regions. The excess mortality rate for people aged 75+ fell significantly in the third wave, partly due to the vaccination schedule and the absence of a normal flu season. For people aged 40–77, the excess mortality rate rose slightly in the third wave. Between regions, excess mortality was highest in Northern Hungary and Western Transdanubia, and much lower in Central Hungary, where the capital is located. The excess mortality rate for men was almost twice as high as that for women in almost all age groups.

## INTRODUCTION

According to aggregated data from official sources, the coronavirus outbreak at the end of 2019 had directly caused the deaths of nearly 5 million people worldwide by September 2021.^2^ The virus emerged in Hungary in March 2020 and by September 2021, the pandemic had significantly transformed our daily lives.^3^ Looking back on this period, one of the important features of the pandemic was that it affected the mortality processes of in Hungary with varying intensity, in terms of time, geography, and age distribution. The literature describes three distinct waves of different viral variants during the period studied: the first was relatively mild in Hungary during the spring of 2020; the second wave, which started in autumn 2020; and the third wave, which started in February 2021.The latter two waves coincided with the winter 2020–2021 influenza epidemic, which was milder than usual due to the protective measures that had been instated against COVID-19 (Frisckle *et* al., 2021; Kung *et* al., 2021). In addition, mortality trends (in a positive direction) were significantly influenced by the vaccination program that started in early 2021 (Vokó *et* al., 2021), which resulted in nearly 60% of the population becoming vaccinated by the end of September 2021.

Our research aims to analyze the mortality trends for the year and a half between March 2020 and September 2021 in terms of the coronavirus. Based on previous years’ age- and gender-specific mortality trends, we estimated how many people would have died in Hungary in the period under study in the absence of the coronavirus pandemic. Then, we compared this estimate with the actual mortality data to obtain the excess mortality. In addition to the number of excess deaths, we calculated the excess mortality rate, which measures the magnitude of excess mortality as a proportion of the relevant population. We used these indicators to describe how the exposure of each age group and the different regions of Hungary changed during successive waves of the pandemic. In addition, we compared the excess mortality with official statistics^4^ on coronavirus victims to show to what extent the other factors (e.g., missed influenza outbreak, vaccination program) affected the overall impact of the coronavirus on mortality.

Many recent publications have described the impact of the pandemic on mortality in Hungary from different perspectives. Some have investigated the spread of the pandemic (Röst *et* al., 2020; Pintér *et* al., 2020) and the effectiveness of vaccines (Vokó *et* al., 2021); others have approached the issue from the perspective of comorbidities and other causes of death (Ostváth *et* al., 2021; Horváth *et* al., 2022), the insurance sector (Csépai and Kovács, 2021), and historical experience (Váradi *et* al., 2020). The geographical spread of the pandemic has also been given particular attention. While some of these studies have compared the domestic experience with that of other countries (Kovalcsik *et* al., 2021), most have focused on geographical differences within countries (Oroszi *et* al., 2021; Uzzoli *et* al., 2021a; Uzzoli *et* al., 2021b).

Several studies have described the evolution of excess mortality in Hungary (Bogos *et* al., 2021; Karlinsky and Kobak, 2021; Túri and Virág, 2021; Páldy and Bobvos. 2021). However, these studies generally compared the Hungarian experience using different dimensions (e.g., period, government measures) with several other countries or groups of countries. Therefore, they calculated only aggregated excess mortality without more detailed breakdowns.

Our study examines excess mortality in Hungary and presents several novel approaches and results compared to those found in other studies. First, we aim to explore in more detail the temporal evolution of excess mortality in the first year and a half of the pandemic in several different disaggregations. In contrast to the studies on excess mortality published by international institutions and research groups which compared countries and categorized populations into five age groups (0–14, 15–64, 65–74, 75–84, and 85+), our study presents national data in five-year age groups for the population aged 35 and over. In addition to the differences between genders and age groups, our research strongly emphasizes presenting regional differences in excess mortality. More detailed data present a more accurate picture of the differences between the waves of the pandemic and help to better understand the background of the difference between excess mortality and official statistics on coronavirus victims.

The first section explains what excess mortality indicates and what this measure should be used for. Then, we review the stochastic mathematical model used to calculate excess mortality, its parameters, and the assumptions used. In the third section, we present the evolution of the national excess mortality for each sex and age group, and the evolution of regional differences. We then compare the trends in excess mortality with official data on coronavirus deaths and analyze the possible reasons for the noted differences. We conclude the paper with a summary.

## 1. WHAT DOES EXCESS MORTALITY MEASURE?

The impact on mortality of an pandemic, virus, or any other event that influences life prospects is often measured by the excess mortality (Collins *et* al., 1930; Collins, 1932). This indicator compares the actual mortality trends with a hypothetical (counterfactual) situation based on the assumption of what would have happened if the event—in this case the coronavirus pandemic—had not occurred. It is important to emphasize that the indicator is an estimate; it requires a forecast of how many people would have died in Hungary if mortality had followed the trends of previous years. This estimation is compared with the actual mortality data to obtain the excess mortality.

One of the two important features of the indicator is that it includes all effects that divert mortality from its historical path (Ackley *et* al., 2021). In the case of the COVID-19 pandemic, it includes both direct and indirect effects. Direct effects include cases where death can be attributed to a coronavirus infection, i.e., someone dies as a direct result of the adverse health effect of the pandemic. The spectrum of positive and negative indirect effects is much broader (Beaney *et* al., 2020). Health system overload, psychological harms associated with the crisis, restrictions on hospital operations that could be postponed, and canceled visits to the doctor all contribute substantially to health risks. However, these negative consequences can be mitigated by increased health financing, stronger protection against influenza due to the general use of masks, and restrictions on more accident-prone (outdoor) activities. The distinction between direct and indirect effects is vague, as it is not always clear whether a death is caused solely by the coronavirus or whether other comorbidities are involved (Tóth, 2021a). This distinction is especially difficult to make in elderly or chronically ill patients. In addition, there is no uniformity in national practice on how to categorize those who are infected but die from other underlying conditions.

Another important feature of excess mortality as an indicator is that, contrary to its name, does not measure the surplus of mortality, but rather the balance in general. The indicator can be negative in cases (such as in Denmark ^5^) where the number of deaths in the period under review is lower than in previous years. For example, there were fewer traffic accidents worldwide during the COVID-19 pandemic and fewer victims of seasonal influenza. Against this background, it is important to stress that excess mortality is different from the official statistics on COVID-19 deaths. The former includes the impact of factors that cannot be directly linked to the adverse health impact of the coronavirus and would presumably not affect mortality trends in the absence of the pandemic.

In Hungary, the most important factor that is indirectly related to the COVID-19 pandemic but significantly influences (reduces) excess mortality is the absence of the seasonal flu. With considerable fluctuations, there were on average 3,000 victims of seasonal influenza in previous winter seasons (Kovács and Pakot, 2020). From the end of 2020 to the beginning of −2021, the flu season in Hungary was practically nonexistent, mainly due to the protective measures introduced against the coronavirus (mandatory mask use, distance control, curfew, etc.). These measures alone saved thousands of lives, reducing excess mortality while leaving the official statistics on COVID-19 deaths unchanged.

While we have only estimates regarding the impact of this abnormal influenza season, in other areas the indirect impact of the pandemic is already reflected in official number of Hungarian Central Statistical Office (HCSO). Detailed exploration of the causal links will require further research, but the breaks in the time series confirm the likelihood that there is a link between the pandemic and the changing number of certain cause of death (Figure 1). As discussed by Osvát *et* al. (2021), after a significant and gradual decline over several decades, the number of suicides rose by 10% last year; conversely, the number of motor vehicle accidents fell by 23% in 2020, after a decade of stagnation.^6^

**Figure 1.:**
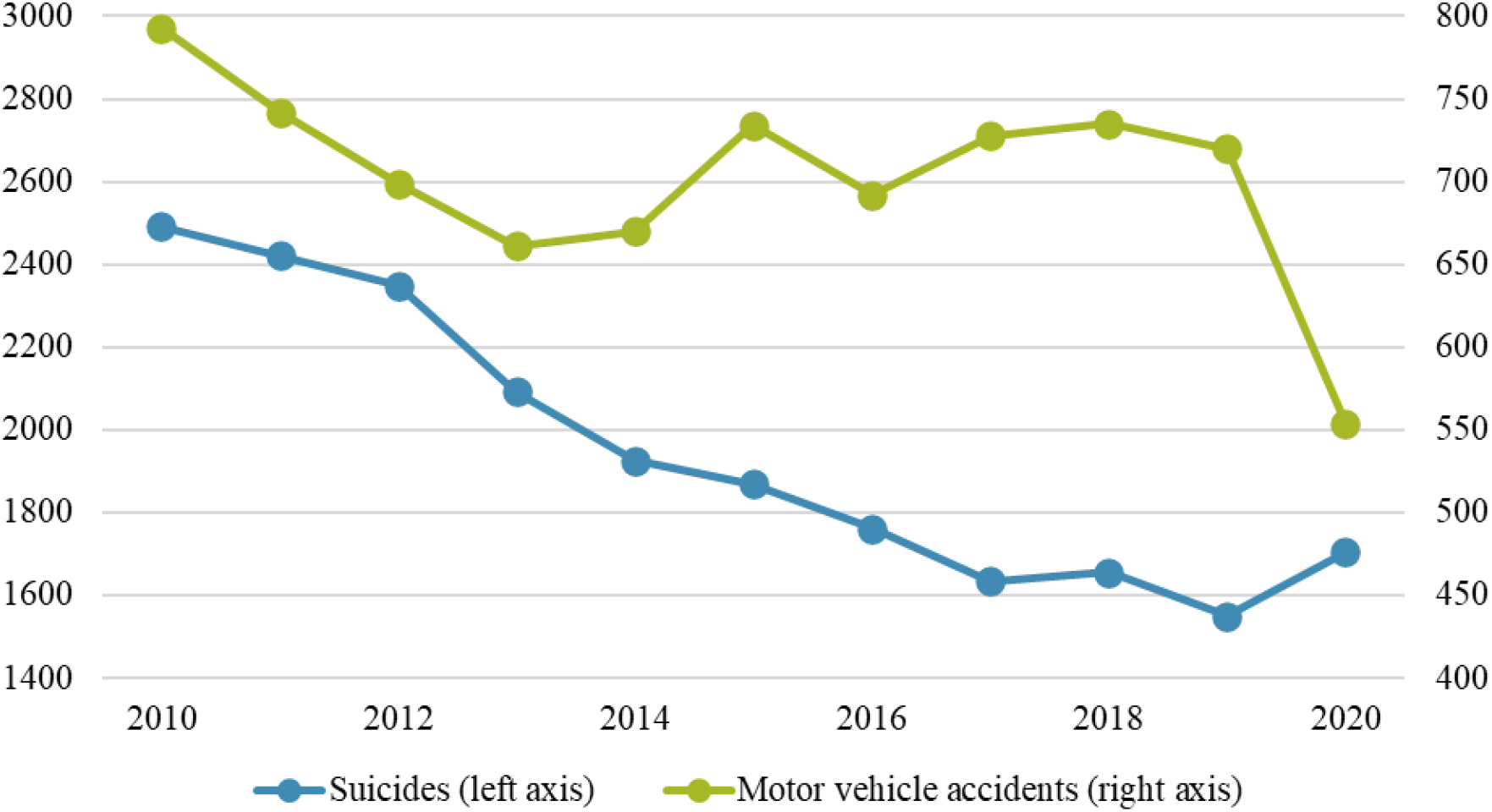
Number of deaths by cause of death. Source: HCSO

While they do not affect the official statistics on COVID-19 deaths, these effects are reflected in the calculation of excess mortality. This is particularly significant because the correlation works in reverse. If we calculate the excess mortality for the last year and a half and compare it with the official statistics on deaths from coronavirus, the difference can be seen as the sum of positive and negative indirect effects.

## 2. DATA AND METHODOLOGY

The Hungarian Central Statistical Office regularly publishes the number of weekly deaths by sex and age group, nationally aggregated and regionally disaggregated. To calculate the excess mortality trend for a given period, we forecasted how many people would have died in the examined period in the absence of the pandemic, following the mortality trends of previous years. For comparability, the structure of the existing data also determined the structure of the forecast. For this reason, it was necessary to estimate both the weekly national mortality rates by sex and age, and the weekly trends in the number of deaths by region. To ensure that our calculations were robust, the two projections were carried out separately, i.e., we produced a projection of the mortality trend both top-down (using aggregated national data) and bottom-up (using regional data). Matching the sum of the regional data with the national data was also as a robustness check.

Weekly forecasting of mortality rates was done in several steps. The first and most important of which was forecasting annual sex- and age-specific mortality rates using a stochastic mathematical model (see also Vanella *et* al., 2021). To achieve this, we used an improved version (Lee-Miller, 2001) of the Lee–Carter (1992) model, which is a classic model for mortality prediction^7^, in both our national and regional studies.^8^

The advent of the Lee–Carter model (1992) began a new era in mortality forecast. Because of its simplicity and accuracy, the model quickly gained worldwide popularity and soon became “the leading statistical mortality model in the demographic literature” (Deaton-Paxson, 2004, p. 264). The strength of this model lies in applying several different time series analysis techniques to historical data to predict mortality. The model predicts the mortality rates for each age group based on the relationship between the mortality rate for that age group and the mortality rate for the whole population over the period. The model aims to explain and predict mortality change by capturing the year effect (longitudinal) and the age effect (cross-sectional). The model is based on the following equation:

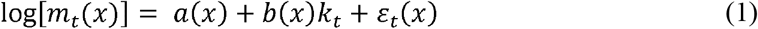

The left-hand side of the equation contains the logarithm of the mortality rate for age *x* in year *t*. The transformation is applied because it precludes the estimated mortality rate from being negative. The right-hand side *a*(*x*) is the average log mortality rate, which represents the average logarithmic mortality rates for each age group and thus represents the typical evolution of mortality by age. Accordingly, the value can reach a minimum shortly after a relatively high value for newborns, and from then it increases with age (Vékás, 2016). Also known as the mortality index, *k*_*t*_ is the only time-dependent component in equation (1) and represents the change in mortality over time. It is usually a decreasing series but often includes short, increasing phases (e.g., during wars). The age-dependent sensitivity *b*(*x*) shows the degree by which the logarithmic mortality rate at a given age increases/decreases when the mortality index (*k*_*t*_) increases/decreases by one unit over a unit of time.

The model includes the (*x*), and *b*(*x*) therefore depends only on age and is constant in time, *k*_*t*_ depends only on time and not age, while *ε*_*x,t*_ depends on age and time. The error term contains effects not explained by the model and is assumed to be independent, with an expected value of 0, identical σ^2^ > 0 with the same variance and a normal distribution. Lee and Carter (1992) introduced two further conditions to ensure that the parameters are unambiguously defined:

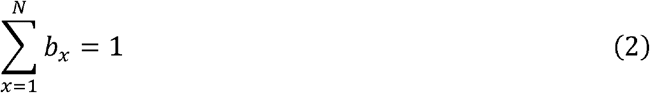

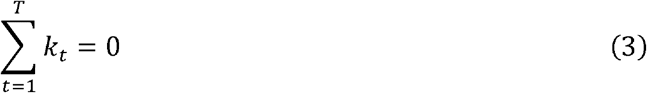

The application of the Lee-Carter model consists of four stages. The first step is to estimate the parameters of the equation using singular value decomposition (SVD), which is the method used to obtain the *M*_*x*_ matrix of mortality data by the least squares to arbitrary precision. In other words, this method decomposes the mortality rate along with the equation (1). In the second step, the resulting *k*_*t*_ parameters are adjusted so that the observed and forecasted mortality rates are the same each year. In the third step, the mortality index is projected into the future. In their original study, Lee and Carter considered the time series of the mortality index an ARIMA process; they found that the model specification of random walk was appropriate based on the data. After predicting the mortality index using the previously obtained average log mortality rate and age-dependent sensitivity, the logarithm of the sex- and age-specific mortality rate for the predicted year can be obtained. Then, the mortality rate is calculated using the equation (1).

The annual mortality rate describes how the number of deaths in an area is proportional to the population living there. The mortality rate must be multiplied by the midyear population to obtain the number of deaths. For both national and the regional level projections, the HCSO has published the sex- and age-specific midyear populations for 2020; for 2021 we calculated the value using a cohort-component method, which takes into consideration the population aging and the mortality experiences of 2020. In the fourth and final step, we converted the annual frequency sex- and age-specific mortality data into weekly frequency data using the within-year distribution of mortality trends in recent years. We obtained the estimated weekly mortality rate by sex and age group for the year and a half under study. The difference between excess mortality and the actual weekly mortality data determined excess mortality.

This method had two important advantages over the simpler and more common practice of measuring weekly deaths against age group mortality data for the previous few years, or using an average of these. First, this method allowed us to consider the continuous improvement in the mortality situation: life expectancy at birth in Hungary, for example, increased from 74.4 years in 2010 to 76.2 years in 2019^9^. Using the sex- and age-specific model was also advantageous because it allowed us to manage the variation in the number of births in different years. This was particularly important for the 65+ age group because of the Ratko generation. For example, between 2019 and 2020, the number of people aged 60–64 years fell from 695,000 to 651,000, a decrease of more than 6%. The improvement in mortality trends and the differences in the number of births between generations are specific features of the demographic trends in Hungary that should be considered when calculating excess mortality.

## 3. RESULTS

According to official data, the first Hungarian victim of the coronavirus pandemic died on 16 March 2020, so we will examine the evolution of excess mortality from week 12 until mid-September 2021 (week 37). During this year and a half, our model calculations suggest that 191,600 people would have died in Hungary without the pandemic, if the mortality trends in Hungary had followed the trends of previous years. In contrast, the weekly data releases of the HCSO indicated that 220,100 people lost their lives in the period studied. Therefore, the excess mortality in Hungary in the year and a half following the pandemic outbreak was 28,400, which represents a 15% increase as compared with the mortality estimated without the pandemic. In most of the studies that quantified excess mortality in Hungary, the investigated period ended earlier, making it difficult to compare our results with the literature. However, the previous results covering these shorter periods are in line with our findings (Karlinsky and Kobak, 2020; Bogos *et* al., 2021; Túri and Virág, 2021; Páldy and Bibvos, 2021). Minor variations can be explained by methodological differences (e.g., influenza treatment, estimation methodology).^10^

To better understand mortality trends, we broke down the investigated period into several periods according to successive waves of the pandemic. The cutoff points were defined as the points in time when a sustained increase in excess mortality began or when the indicator fell below zero (Figure 2). According to this categorization, the year and a half under review can be divided into five periods. In the first and last periods (i.e., from virus emergence to week 35 and from week 29 to week 37 in 2021), there was no significant difference between estimated and actual mortality; the excess mortality associated with the emergence of the COVID-19 was lower than the volatility observed in the mortality trends.

**Figure 2.:**
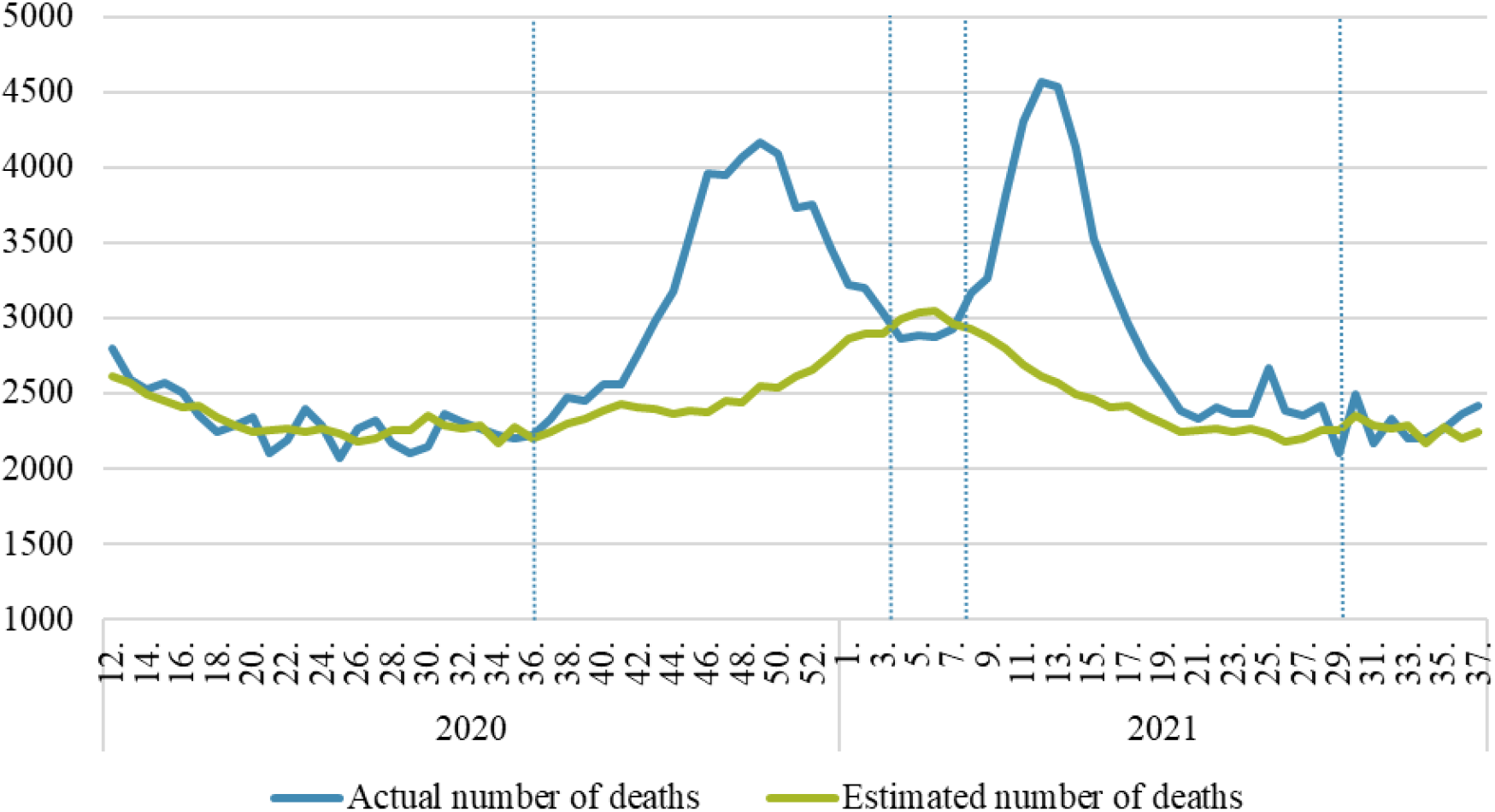
Actual and estimated weekly mortality (persons) Source: own calculations

The second wave of the pandemic reached Hungary in week 36 of 2020 and lasted until week 3 of 2021. Without the pandemic, 52,500 people would have died during this period, whereas 67,700 people actually died. Therefore, during the second wave, the excess mortality in Hungary was 15,200 people, an increase of 29% as compared with the estimated mortality.

Following this wave, the excess mortality was negative for four weeks, meaning that the sum of indirect (negative and positive) effects offset the direct effects. These results can be explained by the observation that the measures introduced to protect against the pandemic (the compulsory wearing of masks, business closures, etc.) also resulted in the containment of the seasonal flu. Given the average of 3,000 deaths from the flu in previous years, we can assume that this also represented a factor of the same magnitude in our mortality projections. The absence of the seasonal flu reduced the excess mortality by approximately the same amount.

The third wave of the pandemic pushed the excess mortality back into the positive range from week 8 onward and continued until week 28. During this period, 51,100 people would have died in Hungary without the pandemic, whereas 64,500 people actually died. In the third wave of the pandemic, the excess mortality was therefore 13,400 people, which was an increase of 26%.

The 18-month trend in excess mortality indicated that the second and third waves of the coronavirus pandemic had a significant impact on domestic mortality trends. While both maintained the indicator in a markedly positive range for 21 weeks, the second wave outperformed the third wave in terms of the number of victims and percentage increase in the number of deaths. However, the weekly peak of nearly 2,000 excess deaths in the third wave was a third higher than the peak of the second wave, suggesting that the impact of the pandemic was more concentrated in the third wave.

### 3.1. AGE-SPECIFIC DIFFERENCES

The excess mortality observed during COVID-19 is represented in different age groups differently: older age groups experience excess mortality more than younger ones in general. Our study investigated how the age pattern was altered by the emergence of new variants, the start of vaccination programs, and other factors such as the absence of the seasonal flu.

First, we observed how the contribution of each group to the total excess mortality evolved. Over the examination period, the share of the excess mortality occupied by people aged 65+ was 77%, 22% for those aged 40–64, and 1% for the younger population. However, the proportion of each age group changed significantly in the third wave as compared with the second; this was most striking for the 75+ age group, whose share of the excess mortality was 60% in the second wave and fell to 37% in the third wave. While the share of the oldest age groups fell significantly, the share of the youngest groups did not show significant change. The share of the excess mortality occupied by those under 40 was 1% in the second wave and rose to 1.5% in the third wave. Therefore, the share occupied by the middle-aged group (aged 40–74) increased substantially from 39% of the total excess mortality in the second wave to 62% in the third wave (Figure 3).

**Figure 3.:**
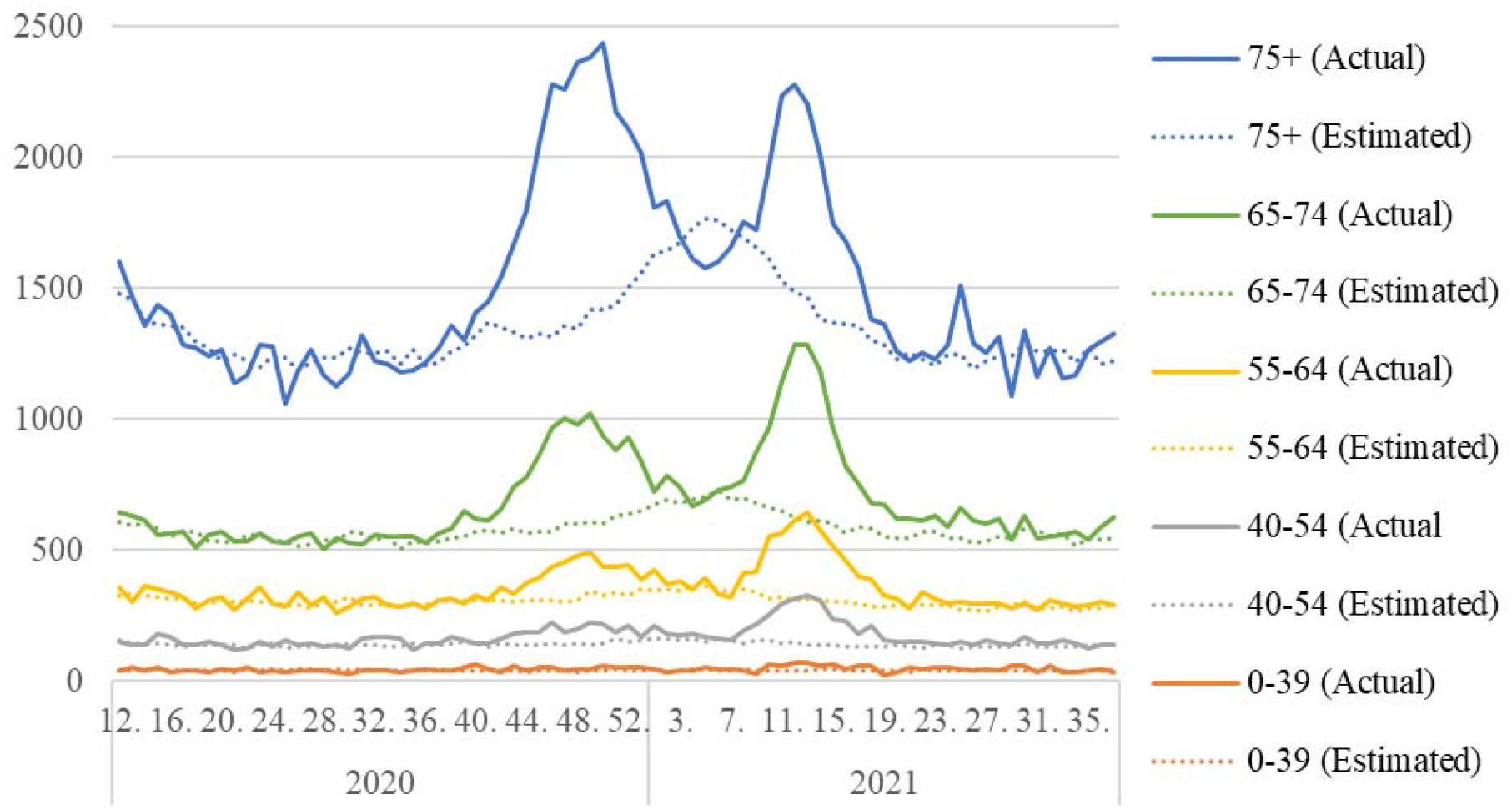
Actual and estimated weekly mortality by age group (persons) Source: own calculations

The decomposition of excess mortality by age group indicates that the second and third waves of the pandemic affected members of different age groups very differently. This phenomenon can be more accurately captured by weighting the excess mortality of the mid-year population of each age group (Figure 4). The excess mortality rate represents the number of excess deaths per 100 persons in each age group. For the oldest age group (85+), this indicator decreased by more than one percentage point in the third wave as compared with the second wave, from 1.9 to 0.6. For the 80–84 age group and the 75–79 age group, the indicator decreased by a smaller degree (0.5 and 0.1 percentage points, respectively).

**Figure 4.:**
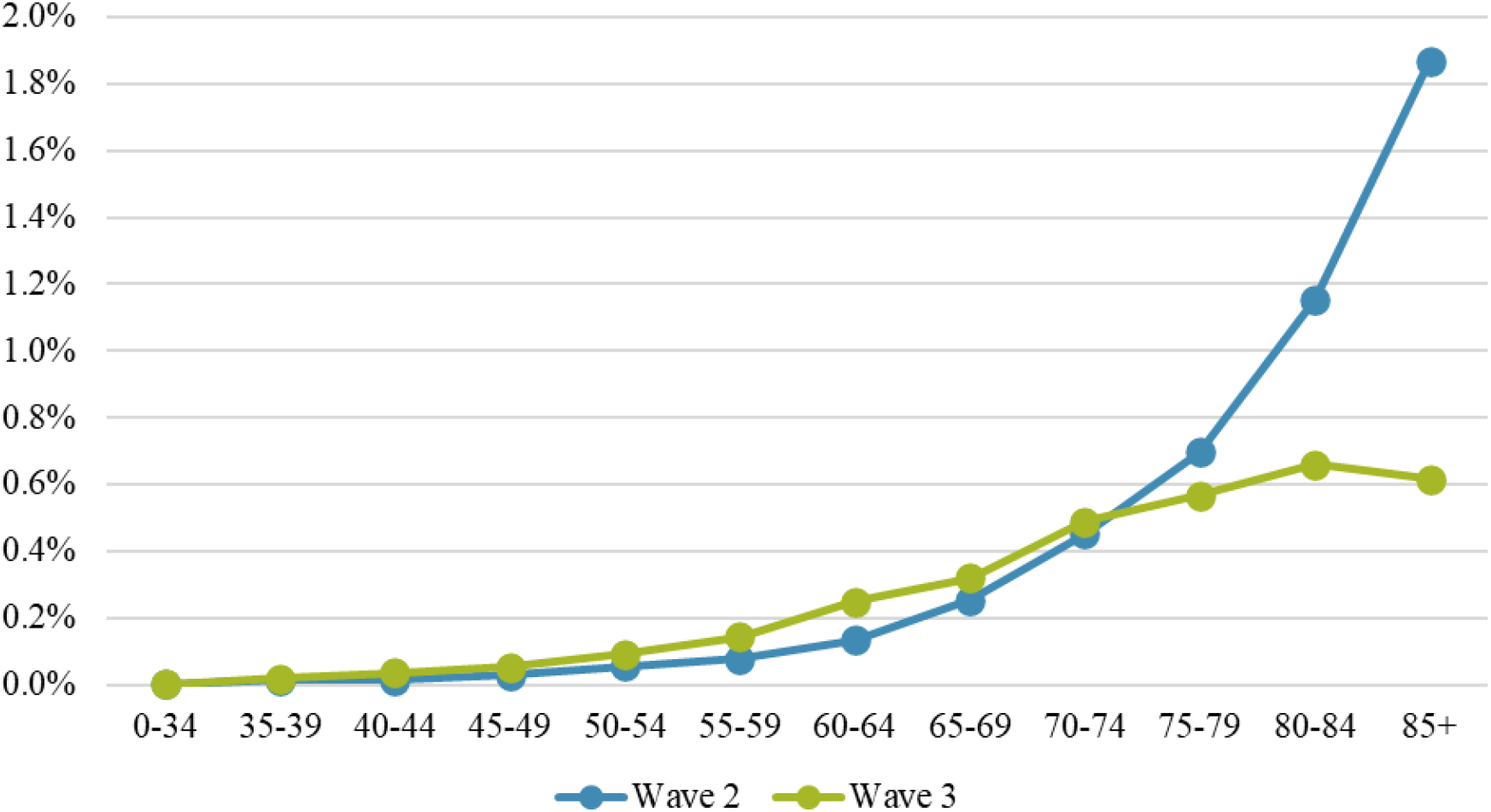
Age-specific excess mortality rate. Source: own calculations

In contrast, the 40–74 age group showed an overall increase of 0.05 percentage points, with the largest increase (0.1 percentage points) in the 60–64 age group. Among those under 40, the excess mortality rate was less than 0.01 percentage points and did not increase significantly. By the third wave, the differences in mortality rates between age groups were significantly reduced. Furthermore, the positive relationship between age and the excess mortality rate was reversed for those over 85, who had a lower excess mortality rate than the 80–84 age group.

The variation in the excess mortality rate for each age group can be explained by several different processes whose detailed exploration is beyond the scope of this paper. However, we will highlight some factors that have contributed to this variation. The most important factor is the emergence of vaccination (Vokó *et* al., 2021). The vaccination program, which started in January of 2021, allowed older people and those in priority professions to enroll first, and gradually opened enrolment to younger age groups. In comparison, the impact of the absence of seasonal flu is likely smaller but not negligible, having saved lives (which implies negative excess mortality) and almost exclusively improving mortality rates for the oldest age group.

In addition, the significant proportion of people in the worst health condition who lost their lives in the second wave may have contributed to the improvement in excess mortality among the elderly by substantially reducing the size of the vulnerable population for the third wave.

### 3.2. GENDER-SPECIFIC EXCESS MORTALITY

In addition to the role of age, less emphasis has been placed on examining differences in excess mortality between women and men. Regarding the number of deaths, no significant difference was apparent: over the year and a half studied, the excess mortality was 13,114 (46.2%) for women and 15,296 (53.8%) for men. However, both sexes were not affected at the same rate, as the proportion of women was much higher than that of men among the elderly population most at risk. Because of the difference in life expectancy at birth among women and men, the proportion of women aged 85+ is 73% (52% of the total population), and therefore much higher in the most vulnerable age groups.

To address the difference in life expectancy, we compared the excess mortality rates of men and women by age group. Our main finding was that when the two waves were considered together, the ratio of excess mortality rates for men to women varied around 2 over all ages; the average of these ratios was also 2. Therefore, men were twice as likely to be at risk of excess mortality over the year and a half studied as compared with women in the same age group.

Our results are in line with the findings of Kontopantelis *et* al. (2020) and Modig *et* al. (2021), as the excess mortality of men exceeded that of women in many countries.^11^ Regarding the difference in excess mortality between sexes, our findings support that there may be some regional pattern to this phenomenon in Europe. Indeed, Islam *et* al. (2021) found that in several countries including Poland, the Czech Republic, Slovakia, and Slovenia, the excess mortality rate for men in 2020 was significantly higher than for women. In contrast, there was little or no difference between the two sexes in several Western European countries, such as Denmark, Norway, Greece, Portugal, and Germany.

This difference was observed in the second and third waves, with greater variation among age groups. For those aged 80+, the a ratio of excess mortality rates for men to women was 1.2 in the second wave, rising to 1.7 in the third wave. Given that foreign studies generally report that women are more likely to refuse vaccination (Zintel *et* al., 2021; Kricorian *et* al., 2021), the increase in the difference between the excess mortality rate of men and women should probably be sought in a different direction.

### 3.3. REGIONAL EXCESS MORTALITY

One of the most important questions to address when protecting against COVID-19 is what factors influence the spread of the pandemic and how these factors change during the different phases of the pandemic. An important starting point is to map the geographical characteristics of the pandemic (Oroszi *et* al., 2021, Uzzoli *et* al., 2021a). In this study, we relied on regional level (NUTS-2) analysis, working with relatively few but large geographical units. On the one hand, we minimized the biasing effect of administrative specificities in cases where the place of death differed from the place of residence. On the other hand, we obtained a more manageable and meaningful distribution of the pandemic intensity within the country than if we had used smaller territorial units. Nevertheless, a more accurate understanding of the spread of the pandemic requires the examination of lower administrative levels (county, sub-county, municipality), as aggregated data may mask important relationships.

As the excess mortality in the year and a half was mainly concentrated in the second and third waves, our regional analysis also focused on these two periods. While almost a quarter of the national excess mortality was attributable to the Central Hungary^12^ region (which includes Budapest and Pest County), considered the most intensely affected region, the population ratio shows the opposite situation. The excess mortality rate per 100,000 population was highest in Northern Hungary (356), with West Transdanubia (324) having a significantly higher excess mortality than the rest of the country. In Southern Transdanubia (311), Central Transdanubia (306), and the Southern Great Plain (304), the intensity of the pandemic was similar but slightly higher than in the Northern Great Plain (290), where the situation was similar to the national average (287). Compared with the rest of the country, the impact of the pandemic on mortality was much lower in the Central Hungary region (227), where the excess mortality was less than 80% of the national average.

We are not aware of any published studies that have calculated excess mortality in Hungary at the regional level. However, studies have examined official statistics on COVID-19 deaths at the geographical level. Among them, the work of Uzzoli *et al*. (2021a) is most related to ours, and although they examined a shorter period, our results corroborate theirs. By examining county and sub-county data, they found that while the pandemic was most intense in Western Transdanubia in terms of infections between March 2020 and February 2021, mortality was highest in Northern Hungary during the same period. Furthermore, Northern Hungary had a higher number of deaths per infected person than the national average at this time. These observations suggest that in Northern Hungary, the health status of the infected population and the lack of adequate health care contributed more to excess mortality and less to the number of infections. At the same time, in Western Transdanubia, the number of infections contributed more to the high excess mortality.^13,14^

As the geographical pattern has changed over time, the difference between regions has decreased significantly between the second and third waves (Figure 7). The difference between the region with the highest and the region with the lowest excess mortality per 100,000 population was 1.9 times in the second wave, while it decreased to 1.3 times in the third wave.

**Figure 5.:**
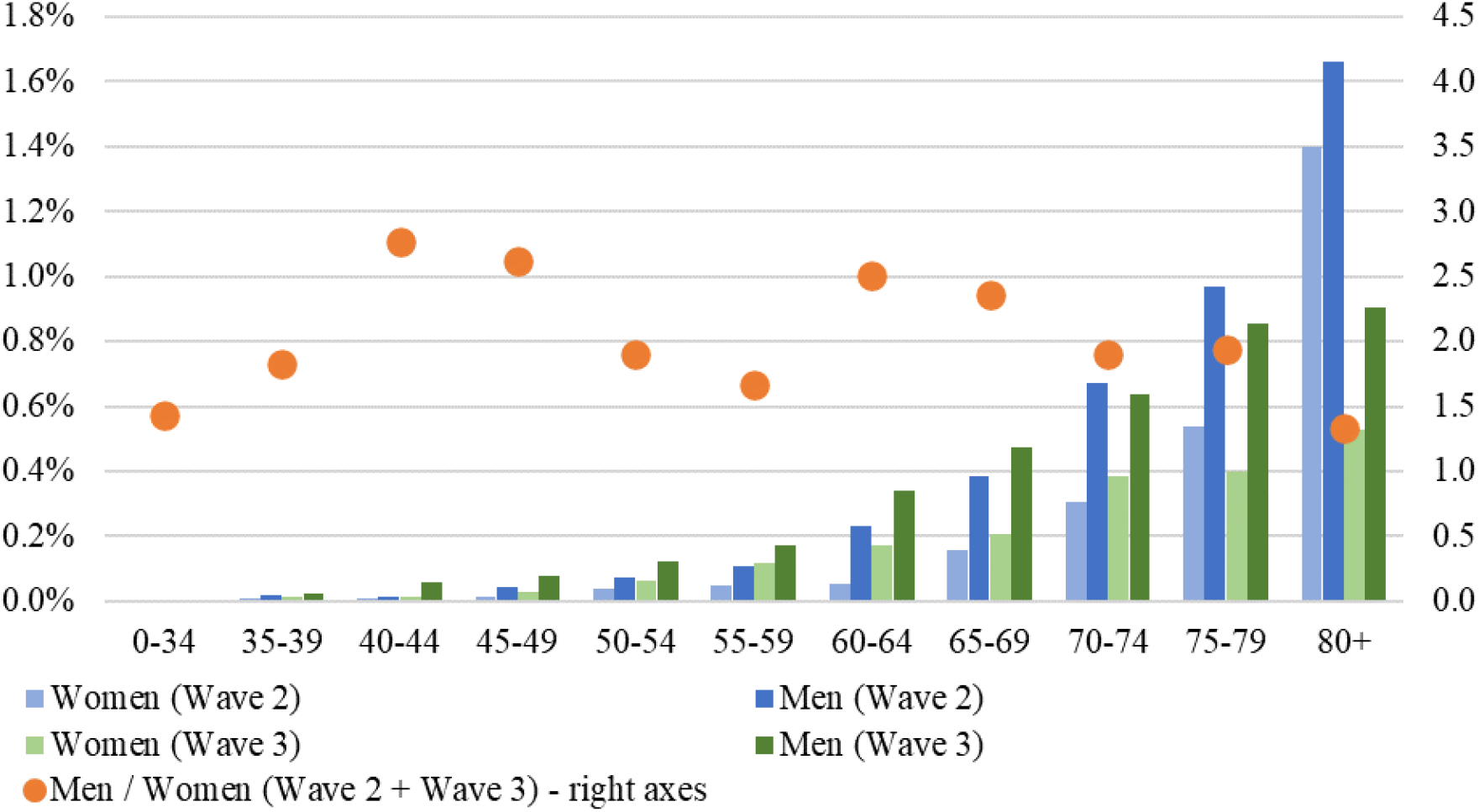
Excess mortality rates by sex and age group in Wave 2 and Wave 3. Source: own calculations

**Figure 6.:**
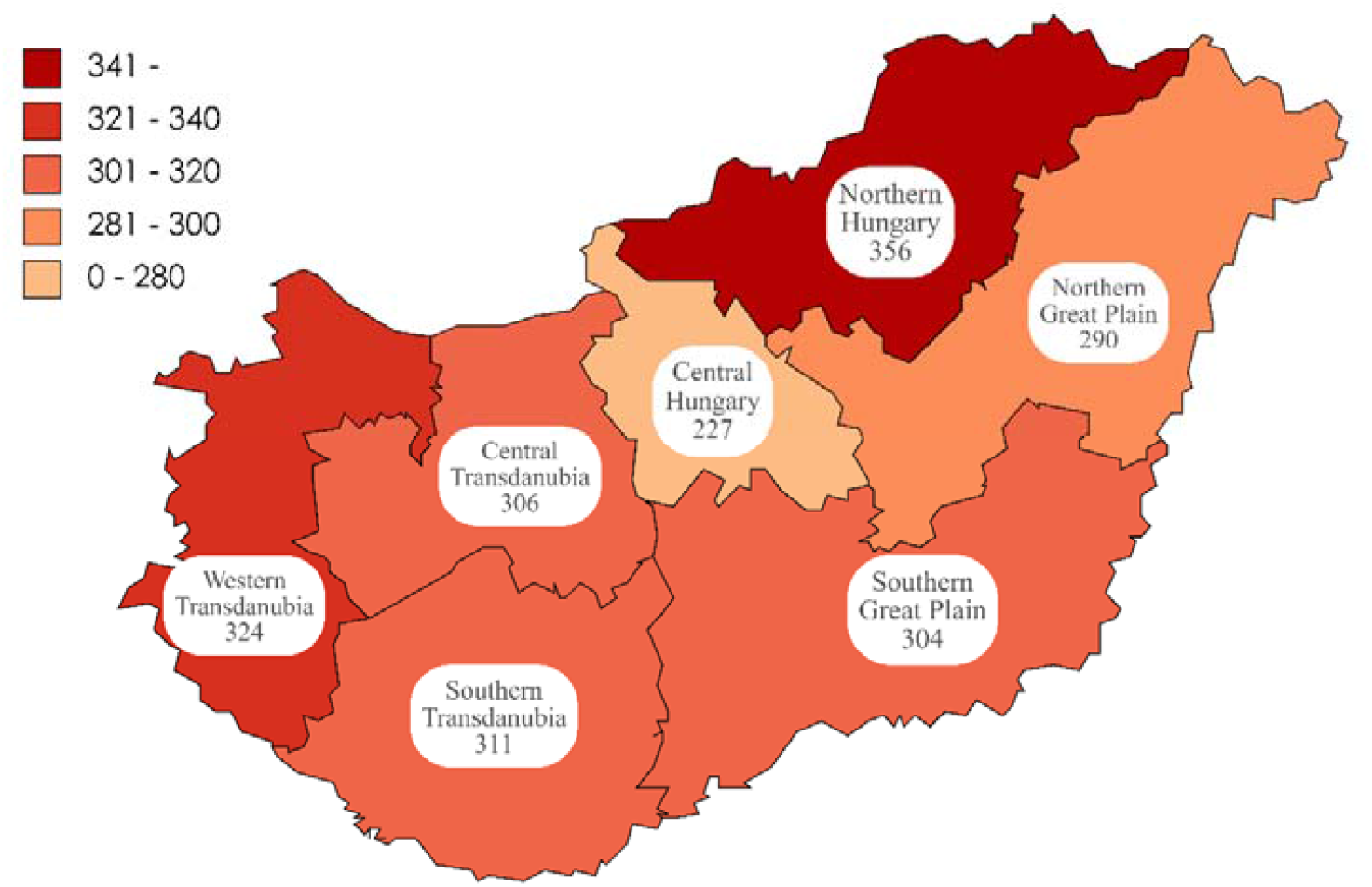
Excess mortality per 100,000 population during Wave 2 and Wave 3, combined. Source: own calculations

**Figure 7.:**
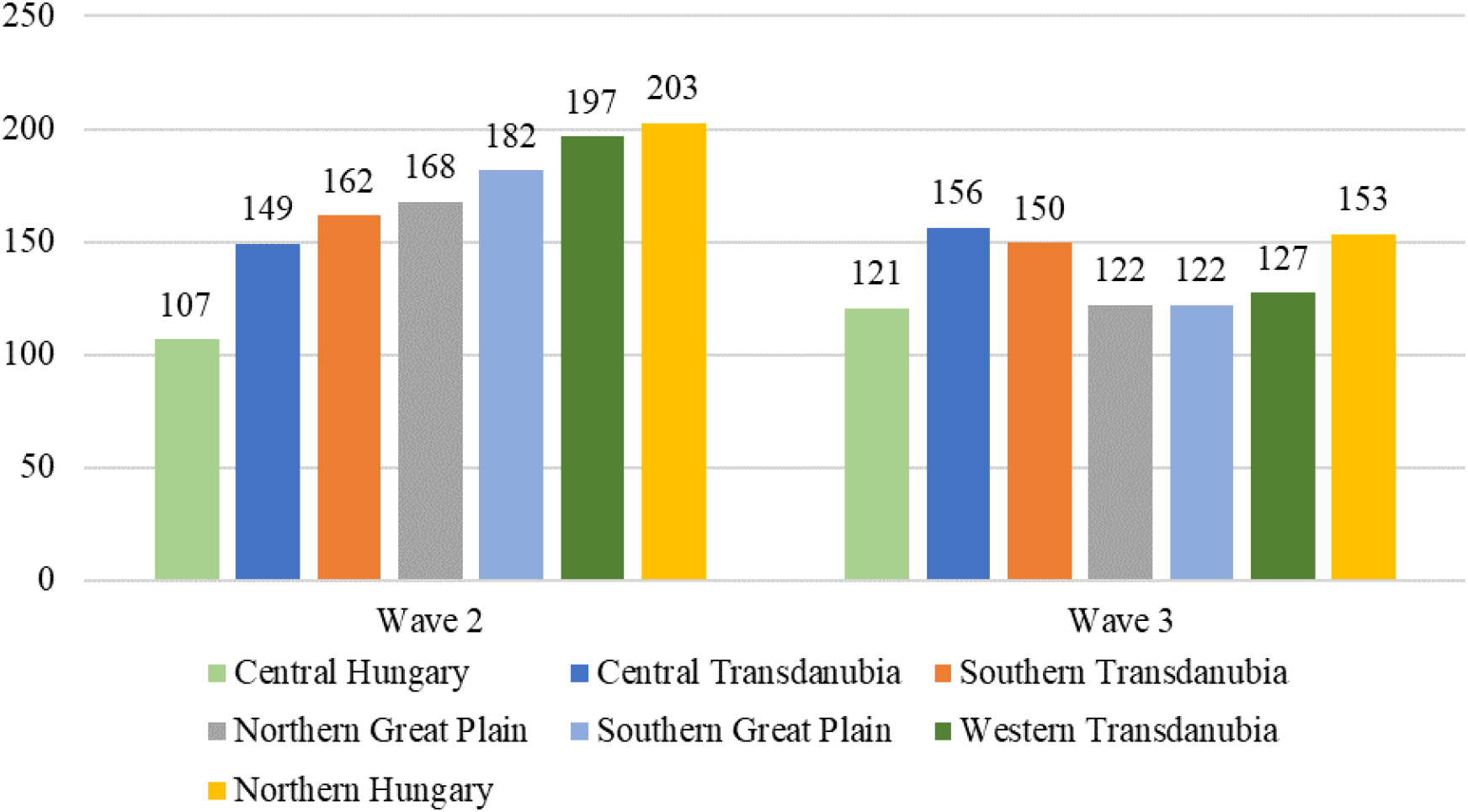
Excess mortality per 100,000 population during Wave 2 and Wave 3. Source: own calculations

In the second wave, however, there were regions with an excess mortality rate of less than 70% of the national average, and others were more than 130% of the national average; in the third wave, there was no region with an excess mortality rate below 90% or above 120% of the national average. These findings suggest that the differences between the second and third waves narrowed considerably at the regional level, which was supported by Kovalcsik *et* al.’s (2021) observation. They studied the changes between the first and second waves in nine Central-European countries and found that regional differences decreased as the duration of the pandemic increased.

### 3.4. EXCESS MORTALITY AND OFFICIAL STATISTICS

Excess mortality is a different measure than the official statistics on COVID-19 deaths; while the latter quantifies the direct impact of the pandemic on mortality, the former also includes indirect positive and negative effects. Several conclusions can be inferred from comparing the two indicators. In most countries, the direct effects of the pandemic on mortality were dominant in the first year and a half. If the excess mortality was significantly higher, especially if it was many times higher than the official statistics on COVID-19 deaths, this may highlight methodological variations in the country’s mortality statistics^15^. Because a death may be caused by several concurrent factors, it is not always clear which factor is the true cause of death. This uncertainty does not generally present a problem if there is adequate transparency in practice. However, in the case of international comparisons, it may become problematic if very different methodologies are used to determine whether a death in a given period was caused directly by the pandemic or not. It is therefore important to compare excess mortality with official statistics on COVID-19 deaths.

The further purpose of this comparison follows a similar approach. The difference between the two statistics helps to map the combined size of the indirect positive and negative impacts. The more time that passes from the outbreak, the greater its significance becomes. Health care overload, delayed surgeries, and missed visits to the doctor can reduce the likelihood of timely diagnosis and appropriate treatment of fatal diseases, indirectly increasing the number of deaths associated with the coronavirus pandemic.

According to official data, from week 12 of 2020 to week 37 of 2021, 30,100 people died of the coronavirus^16^. This is roughly 6% (around 1,700 persons) higher than the excess mortality rate for the same period (28,400 persons). This difference is not significant, however it can initially be regarded as counter-intuitive, indicating that the sum of indirect positive and negative effects has reduced the number of deaths. However, given that the absence of the seasonal flu reduced deaths by roughly 3,000 (based on previous years), which explains why the official statistics on COVID-19 deaths exceed the excess mortality.

In addition to the aggregated data, the evolution of the two indicators over time should be considered, as differences can be observed in this regard (Figure 8). At the beginning of the second wave, the excess mortality started to rise earlier and peaked higher than the official statistics. This rise could be attributable to the reallocation of hospital capacity to manage the pandemic, which may have left many people without care. In addition to the overloading of the health care system, we cannot disregard that administrative problems temporarily hampered the registration of deaths, which may explain why the two indicators began to diverge when they did.

**Figure 8.:**
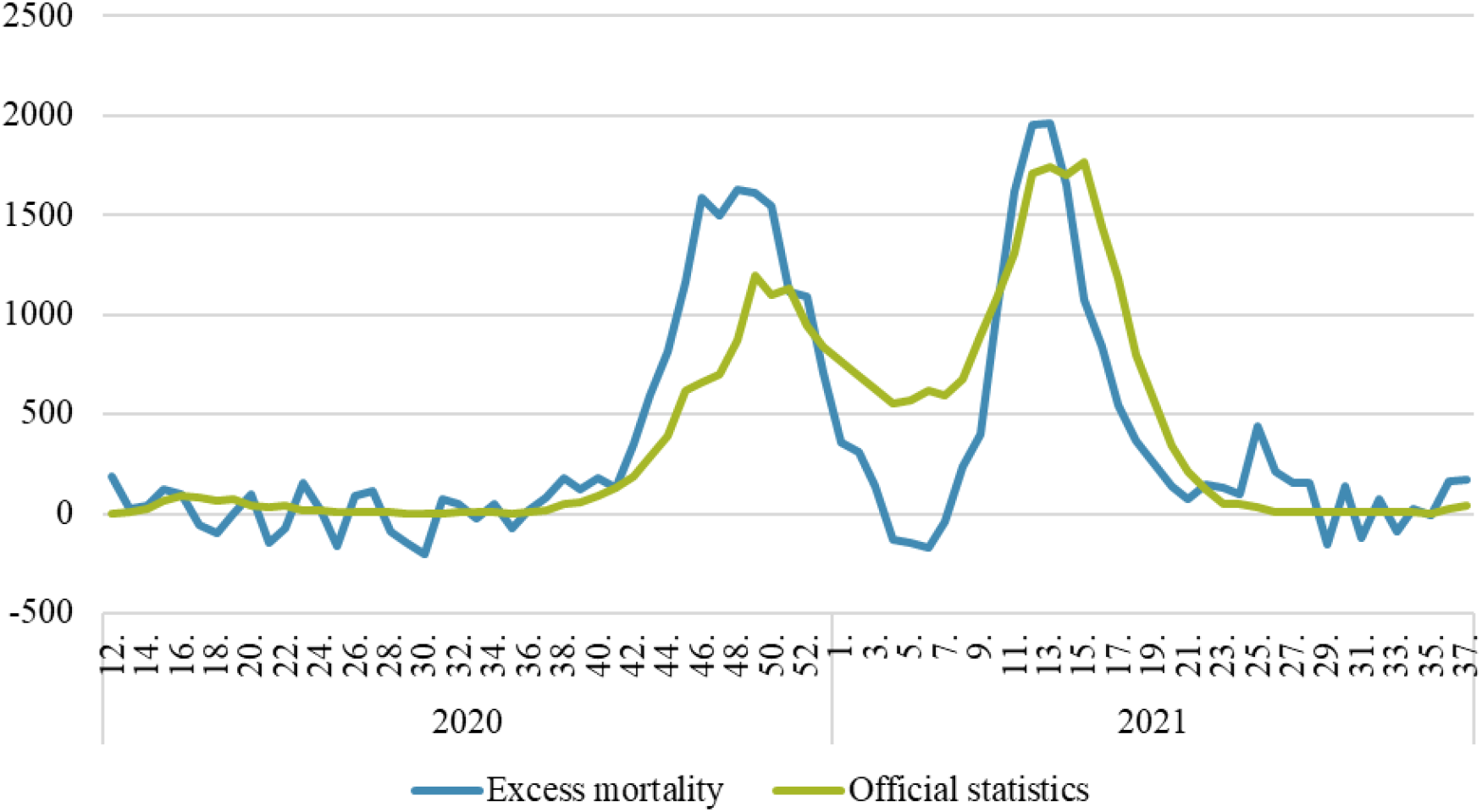
Weekly excess mortality and the official number of COVID-19 deaths (persons) Source: own calculations, koronavirus.gov.hu

The difference between the two waves was also highlighted when official statistics began to show a slight decrease at the first weeks of 2021 in the number of deaths from the coronavirus, but still, more than 500 people died every week. In contrast, the drop in excess mortality at this time was so significant that the indicator remained in the negative range for several weeks. These few weeks coincided with the most intense period of the seasonal flu, and the absence of the flu as an indirect effect reduced the excess mortality but left the official statistics unchanged.

These findings are supported by extending the comparison to age groups. From the 35–40 age group to the 70–74 age group, the two indicators either remained generally the same, or the excess mortality slightly exceeded the official statistics (Figure 9). When we summed up the statistics for the population under 75, we found that the overall excess deaths exceeded the official number of deaths of COVID-19 by 1,200 (9%).

**Figure 9.:**
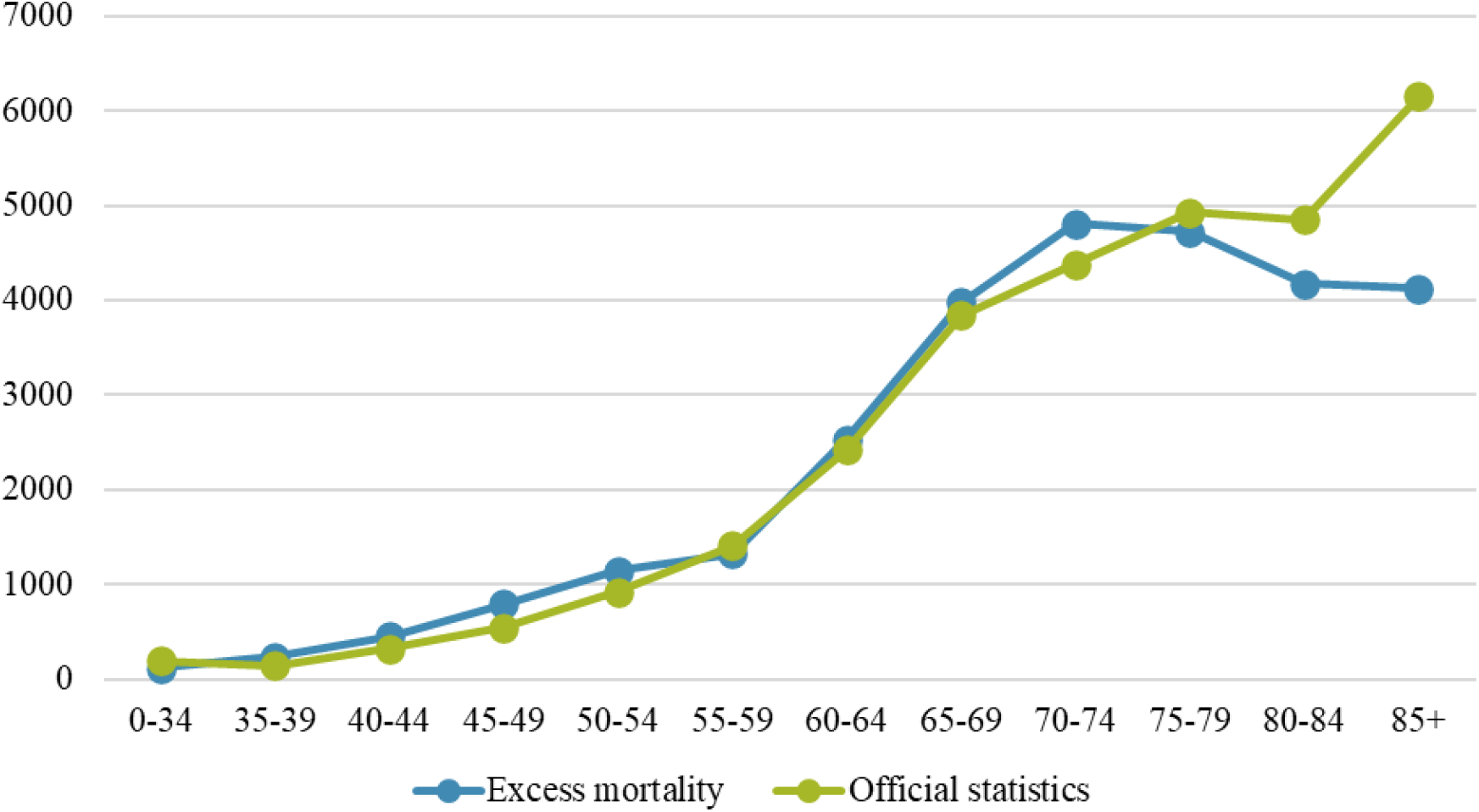
Age-specific excess mortality and the official number of COVID-19 deaths. Source: own calculations, koronavirus.gov.hu ^17^

However, the relationship was reversed for the oldest age groups; the difference remained small for the 75–79 age group, but for the older age groups, the official number of deaths due to COVID-19 was significantly higher than the excess mortality. As the seasonal influenza impacts the oldest age groups the most, the age group breakdown confirms that it is essentially the absence of an influenza pandemic that caused the excess mortality to be slightly below the official statistics.

## CONCLUSION

When examining the impact of an pandemic on mortality, the evolution of excess mortality provides important information. This indicator includes not only deaths directly related to the pandemic but also indirect effects, including factors that increase mortality (e.g., overloaded health services) and reduce mortality (e.g., fewer traffic accidents, absence of seasonal influenza).

COVID-19 reached Hungary in March 2020, and in the following year and a half 15% more people died than would have died in that period without the pandemic. The excess mortality was 28,400 people, which is 1,700 fewer than the official number of deaths due to the COVID-19 pandemic. The lower excess mortality resulted from the absence of a normal flu season, which was a positive consequence of the protective measures against the pandemic (i.e., lockdowns and mask-wearing).

The first wave of the pandemic in Hungary was relatively mild, while the second wave claimed around 15,000 victims and the third wave 13,000. Although the difference in magnitude was not large, there were significant differences between the affected age groups. Partly due to the vaccination program and the absence of a normal flu season, the excess mortality rate of the oldest population fell significantly in the third wave as compared with the second. As a result, the positive relationship between age and the excess mortality rate was reversed in the third wave, with the 85+ age group having a lower excess mortality rate than the 80–84 age group. At the same time, the excess mortality rate increased slightly for those aged 40–74 and did not change significantly for younger people. Therefore, by the third wave, the differences in excess mortality rates between age groups had narrowed. Although the proportion of the excess mortality occupied by men was only slightly higher (54%) over the year and a half studied, the age-specific analysis showed that the excess mortality rate was almost twice as high for men as for women in almost all age groups.

The excess mortality was highest among the geographical regions in Northern Hungary and Western Transdanubia. In contrast, the Central Hungary region, including the capital, had a much lower excess mortality rate than the rest of the country. Our calculations of regional excess mortality indicated that regional disparities decreased significantly in the third wave as compared with the second.

## Data Availability

All data produced in the present study are available upon reasonable request to the authors

## Footnotes

2 Source: www.ourworldindata.com

3 For more details, see Köllő and Reizer (2021), Mohos *et* al. (2020), Kende *et* al. (2021), Sikos *et* al. (2021), Ferenci (2021) and Sulyok *et* al. (2021).

4 https://koronavirus.gov.hu Published by Cabinet Office of the Prime Minister

5 (Karlinsky and Kobak, 2021)

6 https://www.ksh.hu/stadat_files/nep/hu/nep0010.html. Date of download: 28.09.2021.

7 For more information on the models, see Both *et* al. (2006), Booth and Tickle (2008), Vékás (2017), and Tóth (2021b).

8 For the national projection, the data for the 2010–2019 period are taken from the Human Mortality Database, while for the regional projection, the data for the 1980–2019 period are taken from the Hungarian Central Statistical Office.

9 For more on mortality trends in Hungary, see Bálint and Kovács (2021).

10 See Nepomuceno *et* al. (2021) about the sensitivity of excess mortality

11 However, in their study of 27 countries covering the period before the pandemic, Nielsen *et* al (2021) concluded that, regardless of the features of the pandemic, differences in mortality trends between men and women accounted for the difference in excess mortality.

12 Based on the NUTS2 classification for statistical purposes, as of 2018, Budapest and Pest County are two separate regions. However, given that many people living in the agglomeration work in the capital and their movements are particularly important in an epidemiological study, we treated the two regions as one region (Central Hungary), following the classification before 2018.

13 This is in line with the results of Oroszi *et* al. (2021).

14 Although the age structure of communities influences the impact of the pandemic, regional differences in excess mortality cannot be fully explained by the different demographic characteristics of the regions. Although the proportion of people aged 65+ is lowest in the Northern Great Plain, where the excess mortality was relatively low compared with other regions, the proportion of older people in Hungary is highest in the Southern Great Plain and South Transdanubia. In those regions, the excess mortality was not particularly high. For more details, see Obádovics and Tóth (2021).

15 A comparative analysis by Karlinsky and Kobak (2021) shows that the ratio of excess mortality to the official number of deaths from coronavirus is between 0.5 and 1.5 in two-thirds of EU Member States, compared to 4.5 in Russia, 13.1 in Egypt, 14.5 in Belarus, 31.5 in Uzbekistan and 100 in Tajikistan.

16 https://koronavirus.gov.hu/

17 We are grateful to the editorial staff of atlatszo.hu for providing us with the official statistics in detail and in available form to research use.

## REFERENCES

Ackley, C. A., Lundberg, D. J., Ma, L., Elo, I. T., Preston, S. H. and Stokes, A. C. (2021), County-Level Estimates of Excess Mortality Associated with COVID-19 in the United States. MedRxiv, 2021.04.23.21255564. https://doi.org/10.1101/2021.04.23.21255564

Bálint, L., and Kovács, K. (2018), Mortality. In J. Monostori, P. Őri, and Z. Spéder (Eds.), Demographic Portrait of Hungary 2018, Budapest: Hungarian Demographic Research Institute, pp. 151–178. https://demografia.hu/en/publicationsonline/index.php/demographicportrait/article/view/956

Beaney, T., Clarke, J. M., Jain, V., Golestaneh, A. K., Lyons, G., Salman, D. and Majeed, A. (2020), Excess mortality: the gold standard in measuring the impact of COVID-19 worldwide? Journal of the Royal Society of Medicine, 113(9), pp. 329–334. https://doi.org/10.1177/0141076820956802

Bogos, K., Kiss, Z., Kerpel Fronius, A., Temesi, G., Elek, J., Madurka, I., Cselkó, Z., Csányi, P., Abonyi-Tóth, Z., Rokszin, G., Barcza, Z. and Moldvay, J. (2021), Different Trends in Excess Mortality in a Central European Country Compared to Main European Regions in the Year of the COVID-19 Pandemic (2020): a Hungarian Analysis. Pathology and Oncology Research, 27(1609774), 1609774. https://doi.org/10.3389/pore.2021.1609774

Booth, H., Hyndman, R. J., Tickle, L. and de Jong, P. (2006), Lee-Carter mortality forecasting: A multi-country comparison of variants and extensions. Demographic Research, 15(9), pp. 289–310. https://doi.org/10.4054/demres.2006.15.9

Booth, H. and Tickle, L. (2008), Mortality Modelling and Forecasting: a Review of Methods. Annals of Actuarial Science, 3(1–2), pp. 3–43. https://doi.org/10.1017/s1748499500000440

Collins, S. D. (1932), Excess Mortality from Causes Other than Influenza and Pneumonia during Influenza Epidemics. Public Health Reports (1896-1970), 47(46), pp. 2159–2179. https://doi.org/10.2307/4580606

Collins, S. D., Frost, W. H., Gover, M. and Sydenstricker, E. (1930), Mortality from Influenza and Pneumonia in 50 Large Cities of the United States, 1910-1929. Public Health Reports (1896-1970), 45(39), pp. 2277–2383 https://doi.org/10.2307/4579795

Csépai, O. and Kovács, E. (2021), Koronavírus-járvány adatok és biztosítási hatások elemzése. Biztosítás És Kockázat, 8(3–4), pp. 24–43. https://doi.org/10.18530/bk.2021.3-4.24

Deaton, A. and Paxson, C. (2004), Mortality, Income, and Income Inequality over Time in Britain and the United States. In: Wise A.D. (eds.), Perspectives in the Economics of Aging, Chicago Scholarship Online, pp. 247–286).. https://doi.org/10.7208/chicago/9780226903286.003.0007

Ferenci, T. (2021), Different approaches to quantify years of life lost from COVID-19. European Journal of Epidemiology, 36(6), pp. 589–597. https://doi.org/10.1007/s10654-021-00774-0

Fricke, L. M., Glöckner, S., Dreier, M. and Lange, B. (2021), Impact of non-pharmaceutical interventions targeted at COVID-19 pandemic on influenza burden – a systematic review. Journal of Infection, 82(1), pp. 1–35. https://doi.org/10.1016/j.jinf.2020.11.039

Horváth, R. A., Sütő, Z., Cséke, B., Schranz, D., Darnai, G., Kovács, N., Janszky, I. and Janszky, J. (2022), Epilepsy is overrepresented among young people who died from COVID-19: Analysis of nationwide mortality data in Hungary. Seizure, 94, pp. 136–141. https://doi.org/10.1016/j.seizure.2021.11.013

Islam, N., Shkolnikov, V. M., Acosta, R. J., Klimkin, I., Kawachi, I., Irizarry, R. A., Alicandro, G., Khunti, K., Yates, T., Jdanov, D. A., White, M., Lewington, S. and Lacey, B. (2021), Excess deaths associated with covid-19 pandemic in 2020: Age and sex disaggregated time series analysis in 29 high income countries. The BMJ, 373: n1137 https://doi.org/10.1136/bmj.n1137

Karlinsky, A. and Kobak, D. (2021), Tracking excess mortality across countries during the covid-19 pandemic with the world mortality dataset. ELife, 10. https://doi.org/10.7554/eLife.69336

Kende, Á., Messing, V. and Fejes, J. B. (2021), Hátrányos helyzetű tanulók digitális oktatása a koronavírus okozta iskolabezárás idején. Iskolakultúra, 31(2), pp. 76–97. https://doi.org/10.14232/iskkult.2021.02.76

Kontopantelis, E., Mamas, M. A., Deanfield, J., Asaria, M. and Doran, T. (2020), Excess mortality in England and Wales during the first wave of the COVID-19 pandemic. Journal of Epidemiology and Community Health, 75, pp. 213–223. https://doi.org/10.1136/jech-2020-214764

Kovács, K. and Pakot, L. (2020), Influenzához kapcsolódó halálozás 2009/2010 és 2016/2017 között Magyarországon. Orvosi Hetilap, 161(23), 962–970. https://doi.org/10.1556/650.2020.31725

Kovalcsik, T., Boros, L. and Pál, V. (2021), A COVID-19-járvány első két hullámának területisége Közép-Európában. Területi Statisztika, 61(3), pp. 263–290. https://doi.org/10.15196/TS610301

Köllő, J. and Reizer, B. (2021), The impact of the first wave of the COVID-19 pandemic on employment and firm revenues in Hungary. Acta Oeconomica, 71(S1), pp. 93–117. https://doi.org/10.1556/032.2021.00031

Kricorian, K., Civen, R. and Equils, O. (2021), COVID-19 vaccine hesitancy: misinformation and perceptions of vaccine safety. Human Vaccines and Immunotherapeutics. https://doi.org/10.1080/21645515.2021.1950504

Kung, S., Doppen, M., Black, M., Hills, T. and Kearns, N. (2021), Reduced mortality in New Zealand during the COVID-19 pandemic. The Lancet, 397(10268), pp. 25. https://doi.org/10.1016/S0140-6736(20)32647-7

Lee, R. C. and Miller, T. (2001), Evaluating the performance of the Lee-Carter method for forecasting mortality. Demography, 38(4), pp. 537–549. https://doi.org/10.2307/3088317

Lee, R. D. and Carter, L. R. (1992), Modeling and Forecasting U. S. Mortality. Journal of the American Statistical Association, 87(419), pp. 659–671. https://doi.org/10.2307/2290201

Modig, K., Ahlbom, A. and Ebeling, M. (2021), Excess mortality from COVID-19: weekly excess death rates by age and sex for Sweden and its most affected region. European Journal of Public Health, 31(1), pp. 17–22. https://doi.org/10.1093/eurpub/ckaa218

Mohos, A., Mester, L., Barabás, K., Nagyvári, P. and Kelemen, O. (2020), Orvos-beteg kommunikációs gyakorlat szimulált pácienssel a koronavírus-járvány idején. (A COVID-19-pandémia orvosszakmai kérdései). Orvosi Hetilap, 161(33), 1355–1362. https://doi.org/10.1556/650.2020.31930

Nepomuceno, M. R., Klimkin, I., Jdanov, D. A., Alustiza Galarza, A. and Shkolnikov, V. (2021), Sensitivity of excess mortality due to the COVID-19 pandemic to the choice of the mortality index, method, reference period, and the time unit of the death series. MedRxiv, 2021.07.20.21260869. https://doi.org/10.1101/2021.07.20.21260869

Nielsen, J., Nørgaard, S. K., Lanzieri, G., Vestergaard, L. S. and Moelbak, K. (2021), Sex-differences in COVID-19 associated excess mortality is not exceptional for the COVID-19 pandemic. Scientific Reports, 11(1), pp. 1–9. https://doi.org/10.1038/s41598-021-00213-w

Obádovics, C. and Tóth, C. G. (2021), A népesség szerkezete és jövője. In Spéder, Z, Őri, P.and Monostori J. (Eds.), Demográfiai Portré 2021, Budapest: KSH Népességtudományi Kutatóintézet, pp. 251–275.

Oroszi, B., Juhász, A., Nagy, C., Horváth, J. K., McKee, M. and Ádány, R. (2021), Unequal burden of COVID-19 in Hungary: A geographical and socioeconomic analysis of the second wave of the pandemic. In BMJ Global Health, 6:e006427. https://doi.org/10.1136/bmjgh-2021-006427

Osváth, P., Bálint, L., Németh, A., Kapitány, B., Rihmer, Z. and Döme, P. (2021), A magyarországi öngyilkossági halálozás változásai a COVID-19-járvány első évében. Orvosi Hetilap, 162(41), pp. 1631–1636. https://doi.org/10.1556/650.2021.32346

Páldy, A. and Bobvos, J. (2021), Többlethalálozás Európában 2020.10. és 2021.18. hét között az EuroMOMO hálózat alapján. Egészségtudomány, 65(2), pp. 4–18. https://doi.org/10.29179/egtud.2021.2.4-18

Pinter, G., Felde, I., Mosavi, A., Ghamisi, P. and Gloaguen, R. (2020), COVID-19 pandemic prediction for Hungary; A hybrid machine learning approach. Mathematics, 8(6) 890, pp1–20. https://doi.org/10.3390/math8060890

Röst, G., Bartha, F. A., Bogya, N., Boldog, P., Dénes, A., Ferenci, T., J. Horváth, K., Juhász, A., Nagy, C., Tekeli, T., Vizi, Z. and Oroszi, B. (2020), Early phase of the COVID-19 outbreak in hungary and post-lockdown scenarios. Viruses, 12(7), 708, pp. 1–30. https://doi.org/10.3390/v12070708

Sikos, T. T., Papp, V. and Kovács, A. (2021), A hazai vásárlói magatartás változása a COVID-19-járvány első hullámában. Teruleti Statisztika, 61(2), pp. 135–152. https://doi.org/10.15196/TS610201

Sulyok, M., Ferenci, T. and Walker, M. (2021), Google Trends Data and COVID-19 in Europe: Correlations and model enhancement are European wide. Transboundary and Emerging Diseases, 68(4), pp. 2610–2615. https://doi.org/10.1111/tbed.13887

Tóth, C. G. (2021), Többlethalandóság a koronavírus-járvány miatt[Magyarországon 2020-ban. KORFA, 2, pp. 2–5. https://demografia.hu/kiadvanyokonline/index.php/korfa/issue/view/586

Tóth, C. G. (2021), Multi-population models to handle mortality crises in forecasting mortality: A case study from Hungary. Society and Economy, 43(2), pp. 128–146. https://doi.org/10.1556/204.2021.00007

Túri, G. and Virág, A. (2021), Experiences and Lessons Learned from COVID-19 Pandemic Management in South Korea and the V4 Countries. Tropical Medicine and Infectious Disease, 6(4), 201. pp. 1–21. https://doi.org/10.3390/tropicalmed6040201

Uzzoli, A., Kovács, S. Z., Fábián, A., Páger, B. and Szabó, T. (2021), Spatial Analysis of the COVID-19 Pandemic in Hungary: Changing Epidemic Waves in Time and Space. REGION, 8(2), pp. 147–165. https://doi.org/10.18335/region.v8i2.343

Uzzoli, A., Kovács, S. Z., Páger, B. and Szabó, T. (2021), A hazai COVID–19-járványhullámok területi különbségei Szerzők: Területi Statisztika, 61(3), pp. 291–319. https://doi.org/10.15196/TS610302

Vanella, P., Basellini, U. and Lange, B. (2021), Assessing excess mortality in times of pandemics based on principal component analysis of weekly mortality data—the case of COVID-19. Genus, 77(16), pp.1-36 https://doi.org/10.1186/s41118-021-00123-9

Váradi, A., Ferenci, T. and Falus, A. (2020), The coronavirus-induced COVID-19 pandemic: Previous experiences and scientific evidences at the end of March, 2020. Orvosi Hetilap, 161(17), pp. 644–651. https://doi.org/10.1556/650.2020.31830

Vékás, P. (2017), Nyugdíjcélú életjáradékok élettartam-kockázata az általánosított korcsoport-időszak-kohorsz modellkeretben. Statisztikai Szemle, 95(2), pp. 139–165. https://doi.org/10.20311/stat2017.02.hu0139

Vékás, P. (2016), Az élettartam-kockázat modellezése. Budapest: Budapest Corvinus Egyetem. http://unipub.lib.uni-corvinus.hu/4112/

Vokó, Z., Kiss, Z., Surján, G., Surján, O., Barcza, Z., Pályi, B., Formanek-Balku, E., Molnár, G. A., Herczeg, R., Gyenesei, A., Miseta, A., Kollár, L., Wittmann, I., Müller, C. and Kásler, M. (2021), Nationwide effectiveness of five SARS-CoV-2 vaccines in Hungary - The HUN-VE study. Clinical Microbiology and Infection. https://doi.org/10.1016/j.cmi.2021.11.011

Zintel, S., Flock, C., Arbogast, A. L., Forster, A., von Wagner, C. and Sieverding, M. (2021), Gender Differences in the Intention to Get Vaccinated against COVID-19 - a Systematic Review and Meta-Analysis. Available at SSRN 3803323. http://dx.doi.org/10.2139/ssrn.3803323

